# Relationship between Her2 oncoprotein and microvessel density (MVD) in different histological types of gastric adenocarcinoma

**DOI:** 10.1101/2025.08.10.25333385

**Authors:** Gazi Abdus Sadique, Mansurul Islam, Salman Bashar Al Ayub

**Affiliations:** Department of Pathology, Satkhira Medical College, Satkhira; MRCS (UK), Department of Surgery, Shaheed Suhrawardy Medical College, Dhaka; Saturia Upazilla Health Complex, Manikganj

**Keywords:** Cervical carcinoma, Mucin stain, Combined PAS, Alcian Blue

## Abstract

**Background:** Gastric adenocarcinoma is the fifth most common cause of cancer worldwide and sixth most common cancer in Bangladesh. So, it is important to predict the biological behavior of gastric adenocarcinoma to predict the prognosis and select the appropriate treatment. Her2 and MVD correlate with clinical outcomes in many human tumors including gastric adenocarcinoma. Malignant tumors associated with decreased Her2 expression and increased MVD tend to show worse prognosis. The purpose of the present study is to explore the correlation between Her2 amplification and microvessel density in patients with gastric adenocarcinoma.

**Methods:** This cross-sectional study was carried out in the Department of Pathology, Satkhira Medical College, Satkhira, from August 2022 to July 2024, to evaluate the relationship between Her2 and microvessel density in gastric carcinoma. Fifty cases of invasive gastric adenocarcinoma were included in this study. Specimen were processed routinely for Hematoxylin and Eosin stain. Her2 immunostaining and CD31 immunostaining for microvessel were done following standard protocol. Histopathological results were analyzed, and statistical analysis was done using Statistical Package for Social Science version twenty-eight.

**Results:** Among the fifty cases of gastric adenocarcinomas, 15 (30%) were Her2 positive (3+) and 35 (70%) were Her2 negative. Positive Her2 expression was higher in 78.6% of intestinal type of gastric adenocarcinoma whereas diffuse type showed Her2 positivity in 21.4% cases which was not statistically significant (p-value >0.05). Mean MVD in the study group was 17.44 ±9.88. High MVD was found in 18 (58.1%) cases of intestinal type of gastric adenocarcinoma and in only 07 (36.8%) cases of diffuse type of gastric adenocarcinoma. The MVD count in Her2-positive gastric cancer (28.93 ± 10.23) was significantly higher than that in Her2-negative gastric cancer (12.51 ± 3.79), (P < 0.001). 93.3% of Her2 positive cases demonstrated high MVD, in contrast 68.6% Her2 negative cases showed high MVD (P <0.001).

**Conclusion:** These results indicated that intestinal type of gastric adenocarcinoma with positive Her2 expression exhibited higher CD31 positive angiogenesis implying aggressive biological behaviors. So, it would be a reasonable approach to consider Her2 and CD31 simultaneously as target of gastric cancer treatment.

## Background

Gastric cancer is the 4th most commonly diagnosed cancer worldwide and 2^nd^ in common cancer related death. More than 90% of gastric cancers are adenocarcinomas.^1^ Gastric cancer spread to the surrounding important organs at an early stage. Surgical resection is the mainstay of treatment in early-stage cancers, but most patients are diagnosed at an advanced stage which are often unresectable.^2^ The survival rate of patients with advanced resectable gastric cancers, however, remains poor despite chemotherapy. So, early diagnosis of the tumor and institution of molecular targeted therapies can help to prevent recurrence, metastasis and improve patient survival.^3^ More than 90% of gastric cancers are adenocarcinomas. They can be further subdivided into intestinal type and diffuse type of adenocarcinoma.^4^

Her2/neu acts as an oncogene and is related in uncontrolled proliferation of cells.^5^ Amplification of Her2/neu oncogene has become an important biomarker for identifying patients who respond to Her2/neu targeted therapy using humanized monoclonal antibody. A study of Herceptin (Trastuzumab) in combination with chemotherapy compared with chemotherapy alone in Patients with Her2-positive advanced gastric cancer revealed a 26% reduction in the risk of mortality when Trastuzumab was added to the chemotherapy regime for treating advanced gastric carcinoma.^6^ ToGA studies have shown that Her2 positive gastric cancer is a unique subtype of disease requiring different diagnosis and treatment strategies and methods. Many patients with advanced gastric cancer have been benefited from anti Her2 targeted therapy.^7^

Angiogenesis is the process of new capillary vessel formation and plays a central in the tumor growth and metastasis formation.^8^ The numerical value of tumor angiogenesis is defined as microvessel density (MVD). It is measured by immunohistochemical staining using antibodies such as CD31 and others that are specific for vessel endothelium.^9^ Studies have shown that overexpression of Her2 can activate EGFR pathway and in turn promote angiogenesis and promote tumor invasion and metastasis.^10^

The aim of the study was to determine if Her2 overexpression is related to the MVD in gastric adenocarcinoma so that it can have impact on patient selection to both anti Her2 and anti angiogenic therapy.

## Materials and Methods

This cross-sectional descriptive study was carried out in the department of pathology, Satkhira Medical College from August 2022 to July 2024. Approval for the research protocol was obtained from the Ethical Review Committee, Satkhira Medical College, Satkhira prior to the commencement of the study. Routine Hematoxylin and Eosin stain were done in Department of Pathology, Satkhira Medical College. Immunohistochemistry of Her2 and CD31 was done in Bangabandhu Sheikh Mujib Medical University, Dhaka. In this study, 50 histopathologically diagnosed gastric adenocarcinoma were taken as cases. Tissue samples were obtained from gastrectomy and endoscopic biopsies. A pre-tested questionnaire used to collect data from cases.

### Interpretation of Her2 expression

According to the scoring system recommended in 2016 Chinese Her2 Detection Guidelines for gastric cancer,^11^ HER2 antibody was localized to the cell membrane of tumor cells, and the score was 0, 1+, 2+, 3 +. Score 0 indicates no response or <10 % staining of tumor cell membrane. Score 1+ indicated that ≥10 % tumor cells were weak or faintly visible membrane staining, or only part of the cell membrane was stained. Score 2+ indicated that ≥10 % tumor cells had weak to moderate basilar membrane, lateral membrane or complete membrane staining. Score 3+ indicated that >10% of the tumor cells had strong staining of the basal lateral membrane, lateral membrane or complete membrane. Score 0∼1+ was directly judged to be Her2 negative, 2+ and 3+ was directly judged to be Her2 positive.

### Calculation of microvessel density

Weidner’s two - dimensional counting method was used for MVD interpretation.^12^ CD31 staining showed a single stained endothelial cell or a cluster of endothelial cells as a positive microvessel. Firstly, the regions with the most abundant vasculature in tumor margins and tumor center, namely “hot spots” were selected under low magnification, and then 5 fields were randomly counted under high magnification (200 times), with their mean value as MVD count. Any endothelial cell or group of endothelial cells that were stained brown and adjacent to the vessel could be used as a counting microvessel. Vessels with a lumen larger than 8 red blood cells and a thicker wall were not counted.

## Results and observation

50 histopathologically diagnosed gastric adenocarcinoma cases were taken. Patients (case) with age ranged from 30 to 66 years (mean age was 53.34 ± 8.38), 64% (n=32) patients were male, and 36% (n=18) patients were female. Out of 50 gastric adenocarcinoma cases, 31 (62%) cases were intestinal type, and 19 (38%) cases were diffuse type adenocarcinoma. Among the study cases 15 (30%) were Her2 positive (3+) and 35 (70%) were Her2 negative. Positive Her2 expression was found mostly in intestinal type of gastric adenocarcinoma, in 12(78.6%) cases whereas diffuse type showed Her2 positivity in 3(21.4%) cases which was not statistically significant (p-value >0.05). Mean MVD in the study group was 17.44 ± 9.88, with a median value of 13.50. High MVD was found in 18 (58.1%) cases of intestinal type of gastric adenocarcinoma and in only 07 (36.8%) cases of diffuse type of gastric adenocarcinoma. The MVD count in Her2-positive gastric cancer (28.93 ± 10.23) was significantly higher than that in Her2-negative gastric cancer (12.51 ± 3.79), (P < 0.001). 93.3% Her2 positive cases demonstrated high MVD, in contrast 68.6% Her2 negative cases showed high MVD (P<0.001).

**Table 01:** Distribution of the study subjects by their age (n = 50).

| Age (years) | Frequency | (%) |
| --- | --- | --- |
| 30-39 | 4 | 8.0% |
| 40-49 | 8 | 16.0% |
| 50-59 | 27 | 54.0% |
| 60-69 | 11 | 22.0% |
| Mean ± SD | $53.34 \pm 8.38$ | |
| Range (Min-Max) | (30-66) |  |

**Figure 01:**
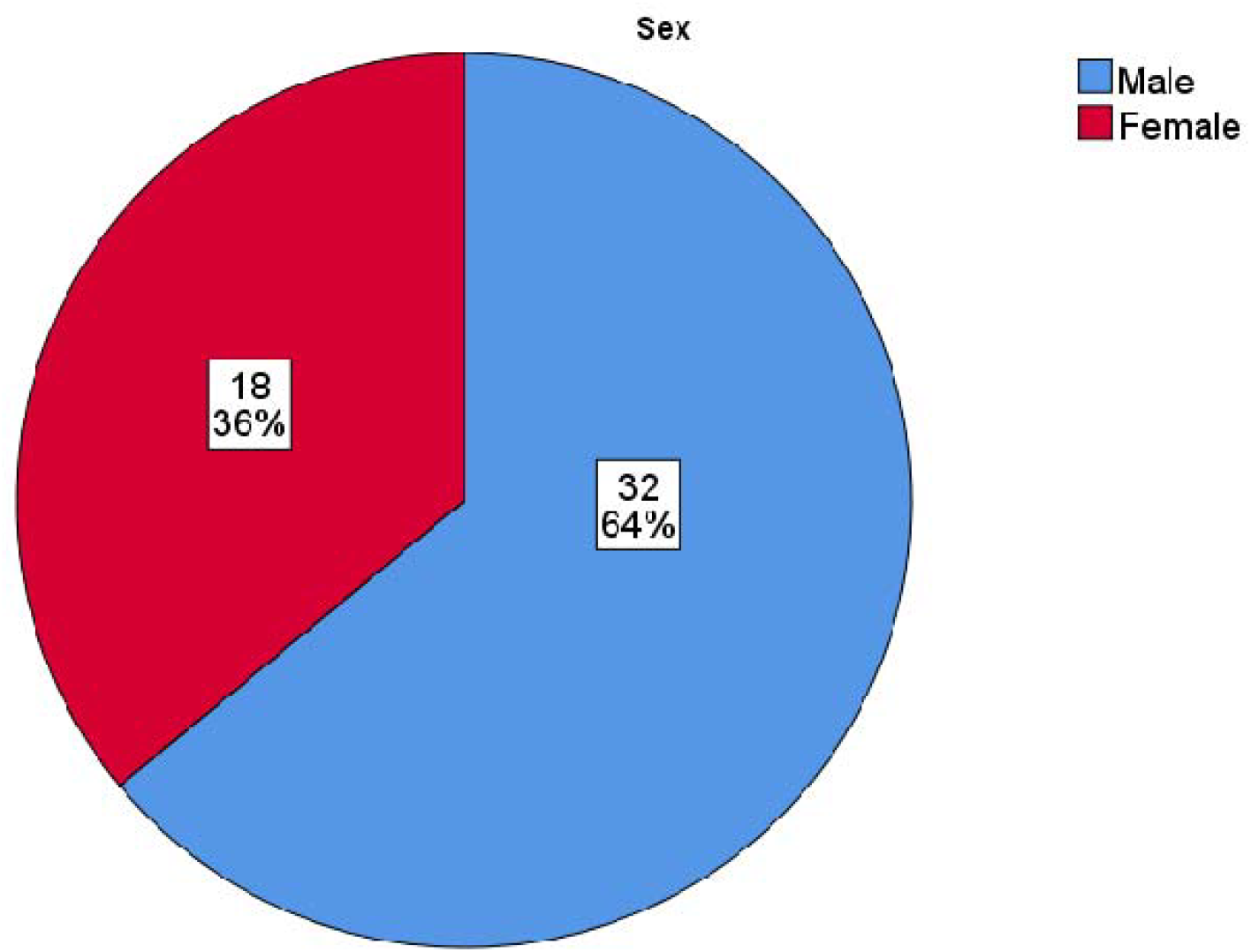
Pie chart showing distribution of cases according to gender (n=50).

**Figure 02:**
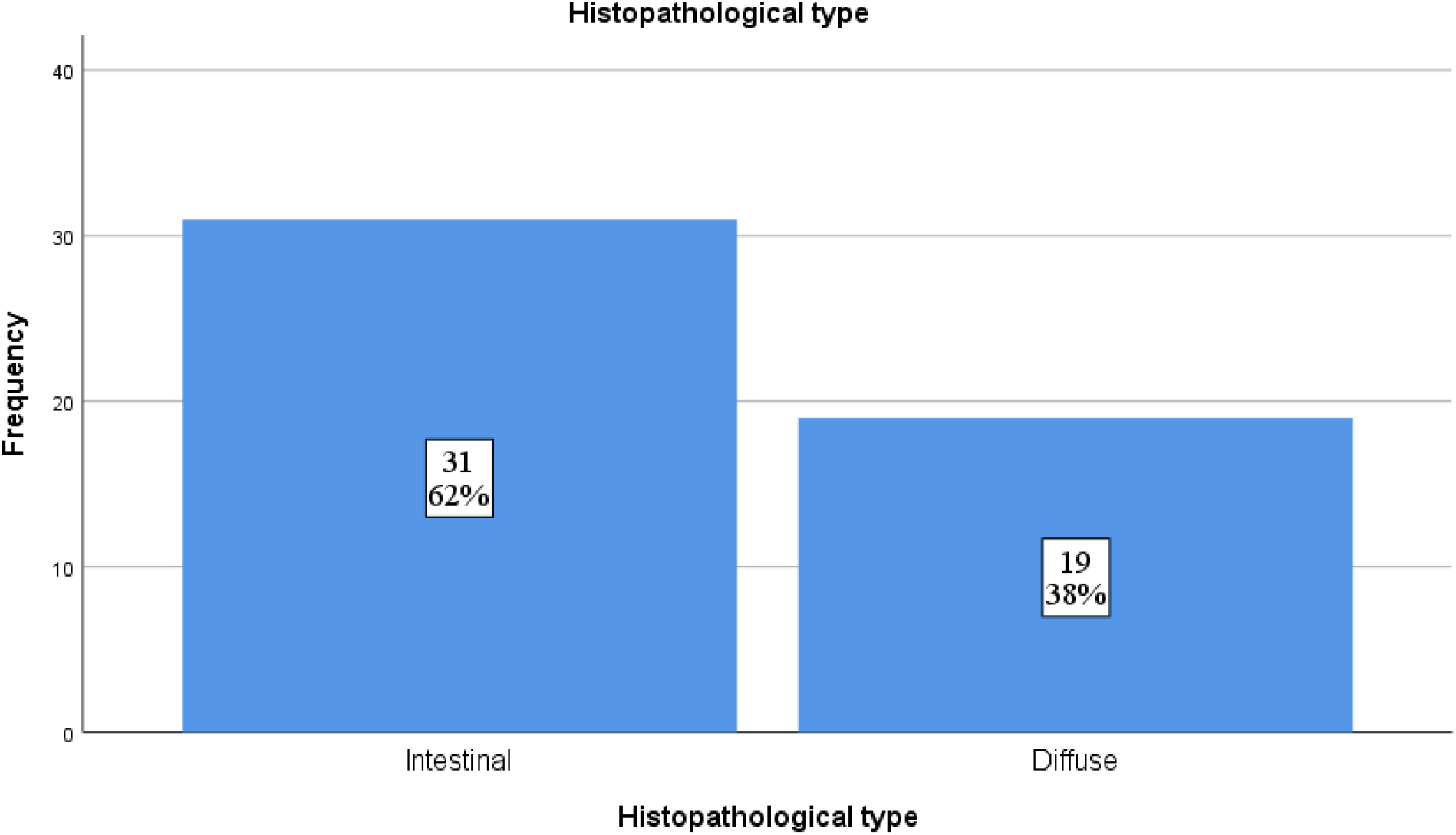
Distribution of tumor type among the study cases (n=50)

**Table 02:** Distribution of the cases according to Her2 expression (n=50).

| Her2 status | Frequency | Percent |
| --- | --- | --- |
| Her2 positive | 15 | 30.0 |
| Her2 negative | 35 | 70.0 |
| Total | 50 | 100.0 |

**Table 03:** Expression of Her2 in different histopathological types of gastric adenocarcinoma (n=50)

| Histopathological type | Intestinal | Diffuse | Total | p-value |
| --- | --- | --- | --- | --- |
| HER2 positive | 12 | 3 | 15 | 0.086 |
|  | 80.0% | 20.0% | 100% |  |
| HER2 negative | 19 | 16 | 35 |  |
|  | 54.3% | 45.7% | 100% |  |
| Total | 31 | 19 | 50 |  |

\*Data were analyzed by using Pearson Chi-Square test.

**Table 04:** Microvessel density among the study groups (n=50).

| Microvessel density | Frequency | Percent |
| --- | --- | --- |
| Low | 25 | 50.0 |
| High | 25 | 50.0 |
| **Mean ± SD** | **17.44 ± 9.88** |  |
| **Median** | **13.50** |  |
| **Range (Min-Max)** | **(6.00-49.00)** |  |

**Table 05:** Association of MVD with histopathological types of gastric adenocarcinoma (n=50).

| MVD status | Histopathological type |  | p-value |
| --- | --- | --- | --- |
|  | Intestinal | Diffuse |  |
| High | 18 | 7 | **0.049** |
|  | 58.1% | 36.8% |  |
| Low | 13 | 12 |  |
|  | 41.9% | 63.1% |  |

\*Data were analyzed using **Pearson Chi-Square Test**.

**Table 06:**
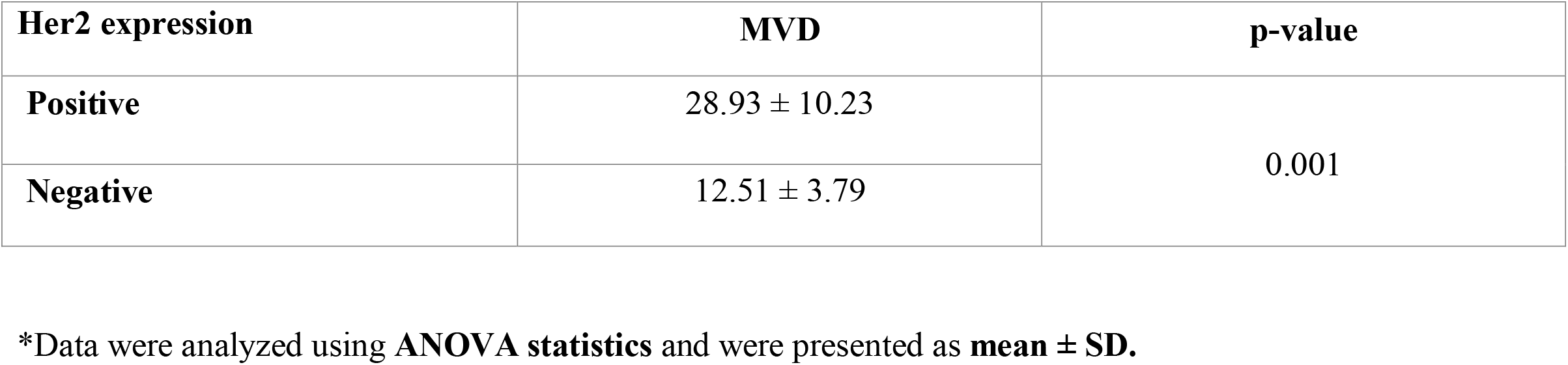
Mean MVD count in terms of Her2 expression (n=50).

| Her2 expression | MVD | p-value |
| --- | --- | --- |
| Positive | 28.93 ± 10.23 | 0.001 |
| Negative | 12.51 ± 3.79 |  |

\*Data were analyzed using **ANOVA statistics** and were presented as **mean ± SD**.

**Table 07:**
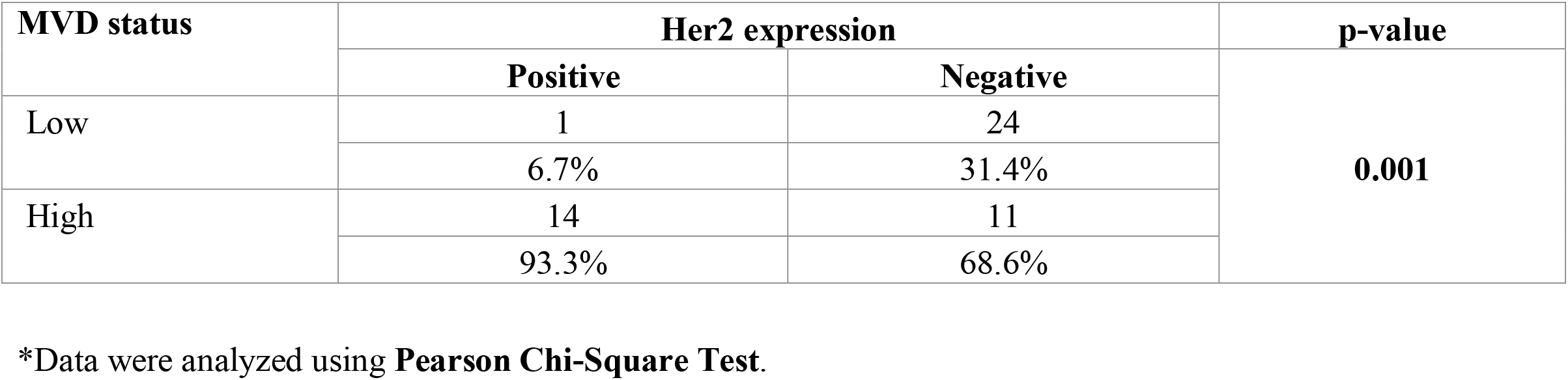
Relationship of MVD with Her2 expression in gastric adenocarcinoma (n=50).

| MVD status | Her2 expression |  | p-value |
| --- | --- | --- | --- |
|  | Positive | Negative |  |
| Low | 1 | 24 | **0.001** |
|  | 6.7% | 31.4% |  |
| High | 14 | 11 |  |
|  | 93.3% | 68.6% |  |

\*Data were analyzed using **Pearson Chi-Square Test**.

**Figure 03:**
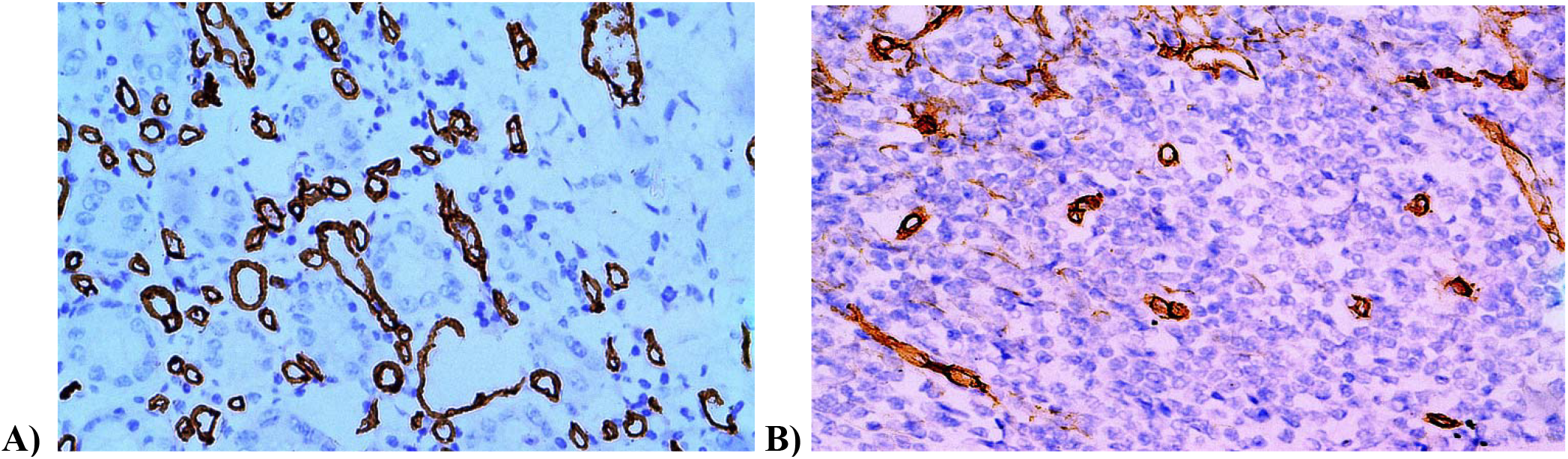
Immunohistochemical CD31 staining was used to mark angiogenesis (MVD). A. Higher MVD counts in intestinal type of gastric adenocarcinoma B. MVD counts were fewer in diffuse type of gastric adenocarcinoma.

**Figure 03:**
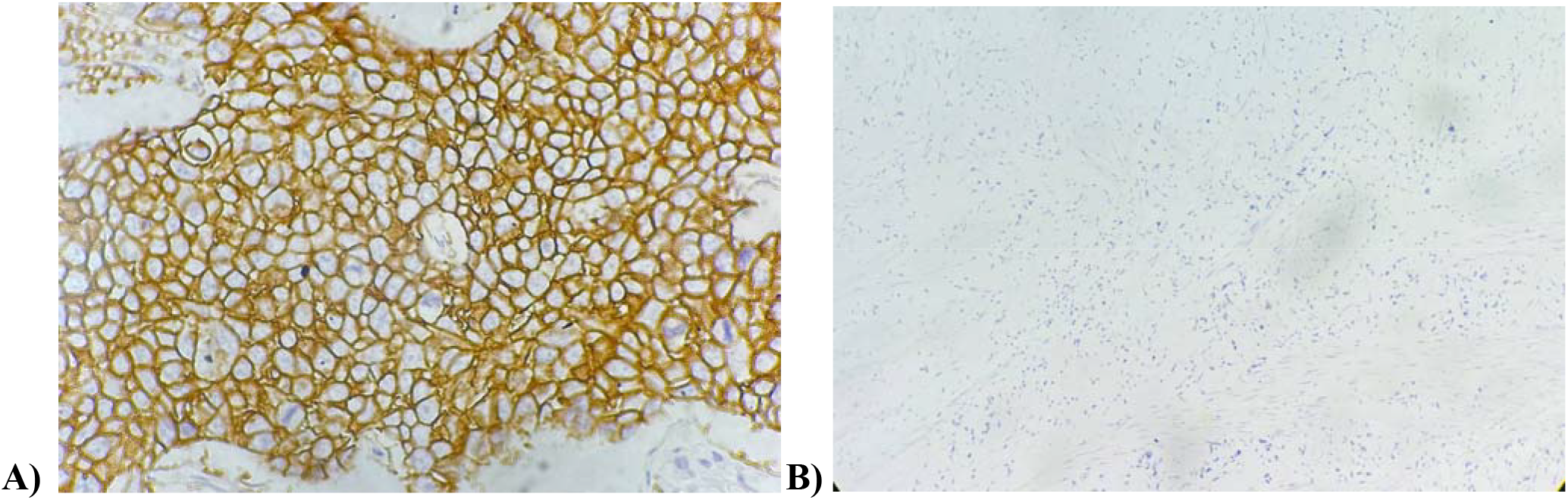
Immunohistochemical staining of Her2. A. Positive Her2 expression in intestinal type of gastric adenocarcinoma B. Negative Her2 expression in diffuse type of gastric adenocarcinoma.

## Discussion

The relationship between Her2 and MVD in gastric cancer is not well established like carcinoma of breast. This has not been widely studied to date. Many authors say that Her2 overexpression implies more aggressive disease course. The precise role of Her2 and MVD in tumor development and progression may be of critical importance for the development and use of new targeted therapies in gastric adenocarcinoma. The objective of the present study was to find out the relationship of Her2 expression and MVD with histopathological types of gastric adenocarcinoma The ages of our studied patients ranged from 30 to 66 years with a mean (53.34 ± 8.38), and we observed that 54% of patients were within 50-59 years of age group. These results are in accordance with another Indian study done by Pramanik *et al*., 2020 where the mean age of gastric carcinoma patients was 55.3, SD 12.71, with an age group ranged from 20 to 81 years.^13^ In our study, 64% of the studied cases were male and 36% were female. Similarly, Abdel-Aziz *et al*, 2017 have reported that among the 48 cases of gastric carcinoma, 30 were male patients (62.5%) and 18 were female patients (29.2%).^14^ In another study conducted by Pramanik et al., 2020 there was a male preponderance (69.2%) with male to female ratio 2.3:1.^13^

Among the present study cases, most were intestinal type 31(62%). Rest was diffuse type 19 (38%). It is consistent with the results of Abel-Aziz et al., 2017 which showed that among 32 cases of gastric adenocarcinoma 66.7% were intestinal subtype and 29.2% were diffuse subtype.^14^ In our study, 15 (30%) were Her2 positive (3+) and 35 (70%) were Her2 negative. It is consistent with the results of Pramanik et al., 2020 where the Her2 positivity was 23.1%. Her2 positive expression was found mostly in intestinal type of gastric adenocarcinoma, in 12(80%) cases and diffuse type showed Her2 positivity in 3(20%) cases which was not statistically significant (p-value >0.05). This result matches that of El-Gendi et al., 2015 which found that 81% of intestinal subtype was HER2 positive and 19% of diffuse subtype was Her2 positive.^15^

In our study cases MVD is 17.44 ± 9.88, with median value 13.50. Badescu et al., 2012 also demonstrated similar findings where the MVD for 28 tumors specimens expressed by CD31 ranged from 12 to 27 with a mean MVD value of 19.14±4.25 SD. Twenty (20) patients were categorized as low MVD CD31 and eight as high MVD CD31.^16^ In our study, high MVD was found in 18 (58.1%) cases of intestinal type of gastric adenocarcinoma and in only 07 (36.8%) cases of diffuse type of gastric adenocarcinoma which closely resembles the findings of Badescu et al. 2012 which stated that MVD value was higher in the diffuse type of gastric cancer in comparison to the intestinal type (33.4 *vs*. 26) (*p*=0.04).^16^

According to our study, the MVD count in Her2-positive gastric cancer (28.93 ± 10.23) was significantly higher than that in Her2-negative gastric cancer (12.51 ± 3.79), (P < 0.001). 93.3% Her2 positive cases demonstrated high MVD, in contrast 68.6% Her2 negative cases showed high MVD (P <0.001). It lies in line with Badescu et al., 2012 where they observed that the mean MVD value is higher in Her2 positive gastric cancer samples than in the negative ones, but this relationship was not statistically significant (*p*=0.29 for CD31 and *p*=0.52 for CD34).

## Conclusion

From the close histopathological and immunohistochemical findings it can be said that Her2 and microvessel density (MVD) in gastric adenocarcinoma showed a positive correlation. The higher the number of MVD, the more expressions of Her2 causing more aggressive tumor progression. This study will help to understand the significance of angiogenesis and growth factor (Her2) in gastric cancer subtypes. These findings may aid the clinicians to select the patients for potential therapeutic targets because concurrent therapeutic strategies against Her2 and angiogenesis may be helpful to control the progression of gastric adenocarcinoma, especially the intestinal subtype.

## Data Availability

All data produced are contained in the manuscript.

## References

1. K.D. Miller, R.L. Siegel, C.C. Lin, A.B. Mariotto, J.L. Kramer, J.H. Rowland, et al., Cancer treatment and survivorship statistics, 2016, CA Cancer J Clin. C.C. (66) (2016) 271–289.

2. Tanner M, Hollmén M, Junttila TT, Kapanen AI, Tommola S, Soini Y, Helin H, Salo J, Joensuu H, Sihvo E, Elenius K, Isola J. Amplification of HER-2 in gastric carcinoma: association with Topoisomerase IIalpha gene amplification, intestinal type, poor prognosis and sensitivity to trastuzumab. Ann Oncol. 2005 Feb;16(2):273–8. doi: 10.1093/annonc/mdi064. PMID: 15668283.

3. Hede K. Gastric cancer: trastuzumab trial results spur search for other targets. J Natl Cancer Inst. 2009 Oct 7;101(19):1306–7. doi: 10.1093/jnci/djp341. Epub 2009 Sep 15. PMID: 19755679.

4. Ma J, Shen H, Kapesa L, Zeng S. Lauren classification and individualized chemotherapy in gastric cancer. Oncol Lett. 2016 May;11(5):2959–2964. doi: 10.3892/ol.2016.4337. Epub 2016 Mar 16. PMID: 27123046; PMCID: PMC4840723.

5. Mrklic I, Bendic A, Kunac N, Bezic J, Forempoher G, Durdov MG, Karaman I, Prusac IK, Pisac VP, Vilovic K, Tomic S. Her-2/neu assessment for gastric carcinoma: validation of scoring system. Hepatogastroenterology. 2012 Jan-Feb;59(113):300–3. doi: 10.5754/hge10776. PMID: 22260838.

6. Moelans, Cathy B., van Diest Paul J., Milne Anya N.A., Offerhaus, G. Johan A., HER-2/neu Testing and Therapy in Gastroesophageal Adenocarcinoma, Pathology Research International, 2011, 674182, 10 pages, 2011. 10.4061/2011/674182

7. Van Cutsem E, Bang YJ, Feng-Yi F, Xu JM, Lee KW, Jiao SC, Chong JL, López-Sanchez RI, Price T, Gladkov O, Stoss O, Hill J, Ng V, Lehle M, Thomas M, Kiermaier A, Rüschoff J. HER2 screening data from ToGA: targeting HER2 in gastric and gastroesophageal junction cancer. Gastric Cancer. 2015 Jul;18(3):476–84. doi: 10.1007/s10120-014-0402-y. Epub 2014 Jul 20. PMID: 25038874; PMCID: PMC4511072.

8. Jomrich G, Schoppmann SF. Targeted therapy in gastric cancer. Eur Surg. 2016;48(5):278–284. doi: 10.1007/s10353-016-0389-1. Epub 2016 Mar 7. PMID: 27795701; PMCID: PMC5065582.

9. Şener E, Şipal S, Gündoğdu C. Comparison of microvessel density with prognostic factors in invasive ductal carcinomas of the breast. Turk Patoloji Derg. 2016;32(3):164–70. doi: 10.5146/tjpath.2016.01366.

10. Ciesielski M, Szajewski M, Pęksa R, Lewandowska MA, Zieliński J, Walczak J, Szefel J, Kruszewski WJ. The relationship between HER2 overexpression and angiogenesis in gastric cancer. Medicine (Baltimore). 2018 Oct;97(42):e12854. doi: 10.1097/MD.0000000000012854. PMID: 30334990; PMCID: PMC6211927.

11. Guideline Recommendations for HER2 Detection in Gastric Cancer Group. [Guidelines for HER2 detection in gastric cancer(2016)]. Zhonghua Bing Li Xue Za Zhi. 2016 Aug 8;45(8):528–32. Chinese. doi: 10.3760/cma.j.issn.0529-5807.2016.08.007. PMID: 27510777.

12. Weidner N, Semple JP, Welch WR, Folkman J. Tumor angiogenesis and metastasis--correlation in invasive breast carcinoma. N Engl J Med. 1991 Jan 3;324(1):1–8. doi: 10.1056/NEJM199101033240101. PMID: 1701519.

13. Pramanik P., Sarker R., Maity M. Study of her2/neu and ki-67 expression in gastric and esophagogastric junction adenocarcinoma and their correlation with grade and stage, International journal of scientific research. 2020. 9(2):2277-8179

14. Abd El-Aziz AM, Ibrahim EA, Abd-Elmoghny A, El-Bassiouny M, Laban ZM, Saad El-Din SA, Shohdy Y. Prognostic Value of Her2/neu Expression in Gastrointestinal Stromal Tumors: Immunohistochemical Study. Cancer Growth Metastasis. 2017 Feb 16;10:1179064417690543. doi: 10.1177/1179064417690543. PMID: 28469470; PMCID: PMC5392021.

15. El-Gendi, Saba & Talaat, Iman & Abdel-hadi. HER-2/Neu Status in Gastric Carcinomas in a Series of Egyptian Patients and Its Relation to Ki-67 Expression. Open journal of pathology. 2015. 5:101–113

16. Bădescu A, Georgescu CV, Vere CC, Crăiţoiu S, Grigore D. Correlations between Her2 oncoprotein, VEGF expression, MVD and clinicopathological parameters in gastric cancer. Rom J Morphol Embryol. 2012;53(4):997-1005. PMID: 23303024.

